## Supplemental material for "Comprehensive analysis of *GBA* using a novel algorithm for Illumina whole-genome sequence data or targeted Nanopore sequencing"

### Supplemental figures

**Figure S1:** Detection of SNVs in homopolymers with ONT.

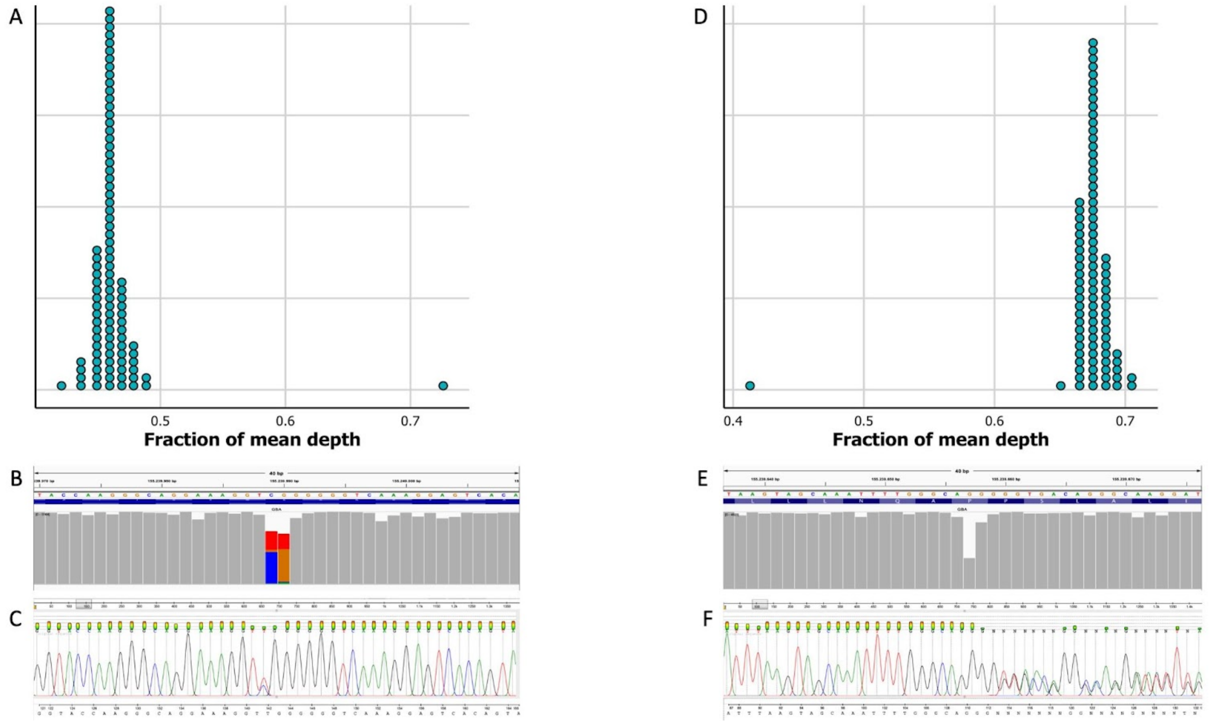

At position chr1:155239990 one sample shows an adjusted depth of coverage that is more than 5 median absolute deviations (MAD) higher than the mean of the other samples **(A)**. Inspection of the sequencing data obtained from ONT (primer pair 1) shows that this is probably due to the presence of a single base substitution at position chr1:155239989 **(B)**, as confirmed by sanger sequencing **(C)**. Similarly, at position chr1:155239657, one sample shows an adjusted depth of coverage that is more than 5 MAD lower than the mean of the other samples, suggesting the deletion of a G in the poly-G. This corresponds to the variant NM_001005742.3:c.413delC **(D)**. Inspection of the sequencing data obtained from ONT (primer pair 1) confirms the drop in depth of coverage **(E)** and Sanger sequencing confirms the presence of a deletion in the poly-G sequence, corresponding to the variant p.P68= **(F)**.

**Figure S2:** Visual display of two reciprocal fusion recombinant alleles.

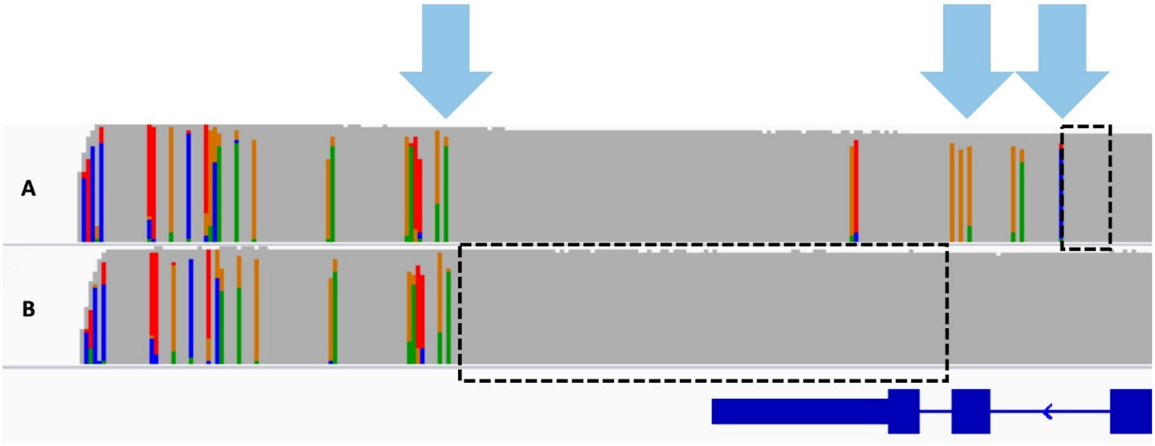

The light blue arrows represent the sentinel positions where *GBA* and *GBAP1* are not homologous. The dotted boxes represent the predicted position of the breakpoints. **A)** breakpoint is between exon 9 and intron 9, so this recombinant allele is predicted to be pathogenic. This is a case of GD, where a pathogenic allele is expected, and indeed had been previously reported independently. **B)** Breakpoint does not affect coding sequence. Exon 11 of *GBA* is 100% homologous to *GBAP1*, so recombinations involving this area do not alter the ORF, so this recombinant is non-pathogenic.

**Figure S3:** Confirmation of the structure of 4 reciprocal recombinant alleles with ONT.

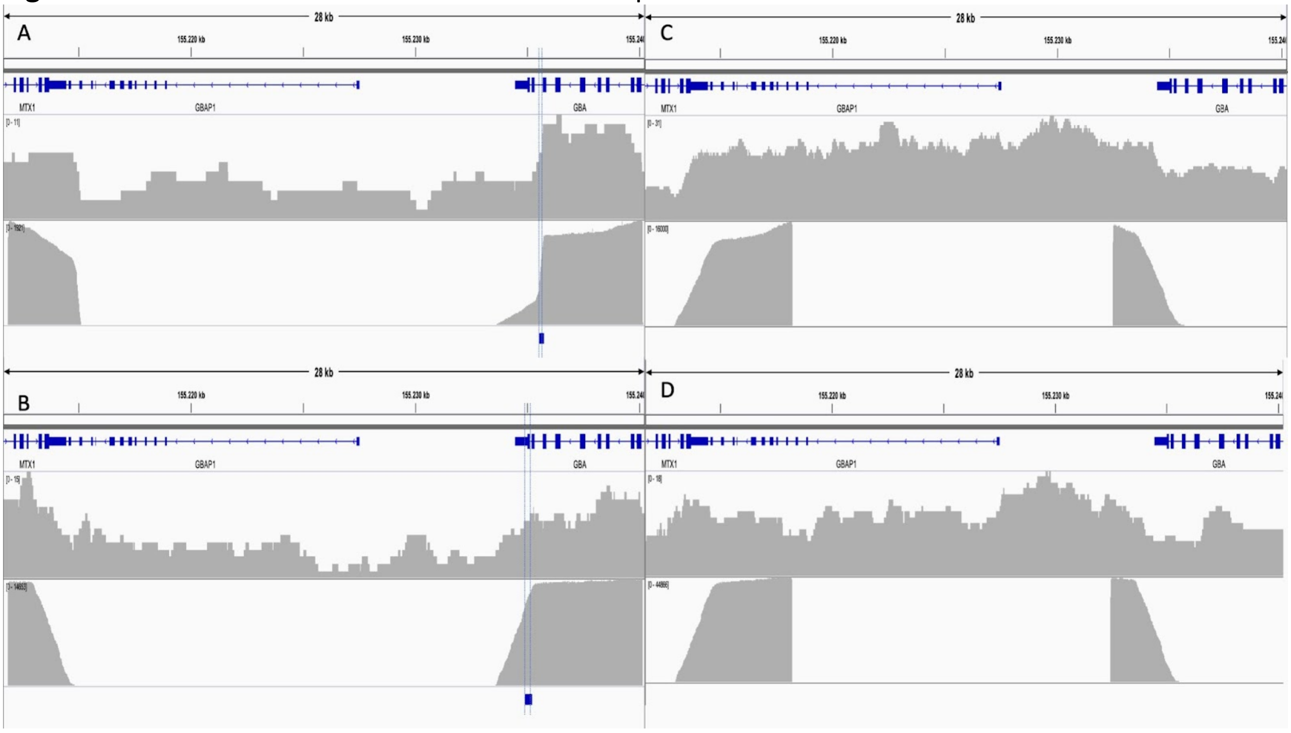

Each panel shows results of one sample: genomic sequencing with adaptive sampling (top), and sequencing of PCR product obtained with primers 2 or 3 (bottom). **A)** Pathogenic CNL allele. **B)** non-pathogenic CNL allele. **C)** CNG allele with a copy number of 5 copies as called by Gauchian 5. **D)** CNG allele with a copy number of 3 copies as predicted by Gauchian.

For the fusion recombinants (A and B), the blue squares and dotted lines at the bottom show the position of the breakpoint predicted with ONT (as described in methods). The breakpoint cannot be refined further as the sequences are identical; see also figure S2. Alignment with LAST.

**Figure S4:** IGV screenshot showing the p.L483R variant.

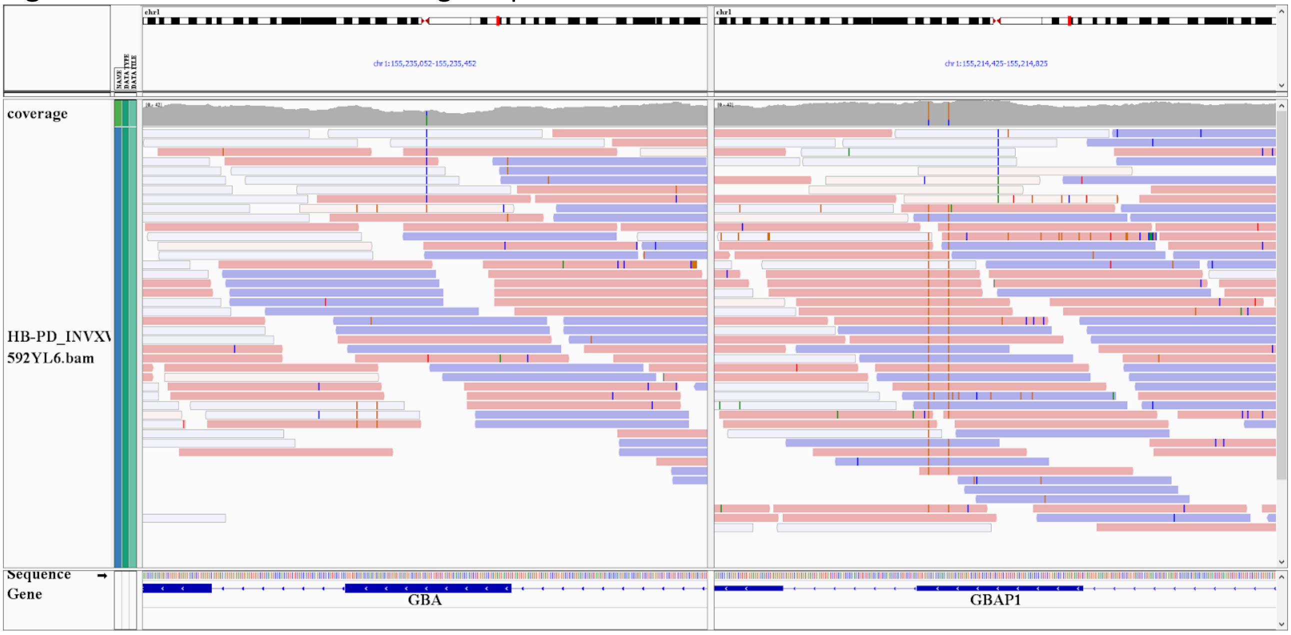

Reads carrying the C variant in the middle are aligned to either *GBA* or *GBAP1*. Many of them have a mapping quality of zero. Hence the variant is missed by BWA-GATK.

### Supplemental tables

**Table S1:** *GBA*/*GBAP1* differentiating sites. Coordinates are in hg38.

| chr | *GBA* position (chr1) | *GBA* base | *GBAP1* position | *GBAP1* base |
|---|---|---|---|---|
| chr1 | 155231496 | c | 155210868 | g |
| chr1 | 155231522 | a | 155210894 | g |
| chr1 | 155231557 | t | 155210929 | c |
| chr1 | 155231692 | g | 155211064 | a |
| chr1 | 155231693 | c | 155211065 | t |
| chr1 | 155231701 | t | 155211073 | c |
| chr1 | 155231818 | aca | 155211190 | a |
| chr1 | 155231834 | g | 155211204 | a |
| chr1 | 155231848 | a | 155211218 | c |
| chr1 | 155231857 | t | 155211227 | c |
| chr1 | 155231870 | a | 155211240 | g |
| chr1 | 155231923 | t | 155211295 | a |
| chr1 | 155231936 | a | 155211308 | g |
| chr1 | 155231937 | g | 155211309 | a |
| chr1 | 155231941 | a | 155211313 | g |
| chr1 | 155231942 | g | 155211314 | a |
| chr1 | 155231989 | g | 155211363 | a |
| chr1 | 155232135 | t | 155211509 | a |
| chr1 | 155232231 | g | 155211605 | a |
| chr1 | 155232340 | c | 155211714 | t |
| chr1 | 155232418 | a | 155211792 | g |
| chr1 | 155232440 | t | 155211814 | c |
| chr1 | 155232447 | a | 155211821 | g |
| chr1 | 155232451 | a | 155211825 | g |
| chr1 | 155232540 | g | 155211914 | c |
| chr1 | 155232549 | c | 155211923 | g |
| chr1 | 155232550 | g | 155211924 | a |
| chr1 | 155232572 | t | 155211946 | c |
| chr1 | 155232721 | c | 155212095 | t |
| chr1 | 155232724 | c | 155212098 | t |

| | | | | |
|---|---|---|---|---|
| chr1 | 155232784 | a | 155212158 | g |
| chr1 | 155232834 | a | 155212208 | c |
| chr1 | 155232892 | g | 155212266 | t |
| chr1 | 155232893 | a | 155212267 | t |
| chr1 | 155232916 | g | 155212290 | c |
| chr1 | 155232919 | a | 155212293 | g |
| chr1 | 155232927 | g | 155212301 | a |
| chr1 | 155232935 | a | 155212309 | g |
| chr1 | 155232982 | g | 155212356 | a |
| chr1 | 155233046 | a | 155212420 | g |
| chr1 | 155233268 | c | 155212642 | g |
| chr1 | 155233287 | g | 155212661 | a |
| chr1 | 155233514 | a | 155212888 | g |
| chr1 | 155233517 | c | 155212891 | t |
| chr1 | 155233521 | t | 155212895 | a |
| chr1 | 155233531 | g | 155212905 | a |
| chr1 | 155233612 | a | 155212985 | g |
| chr1 | 155233639 | g | 155213012 | a |
| chr1 | 155234903 | c | 155214276 | t |
| chr1 | 155235203 | c | 155214576 | g |
| chr1 | 155235217 | c | 155214590 | g |
| chr1 | 155235252 | a | 155214625 | g |
| chr1 | 155235379 | a | 155214752 | g |
| chr1 | 155235412 | g | 155214785 | a |
| chr1 | 155235727 | c | 155215100 | g |
| chr1 | 155235749 | GGGACTGTCGACAAAGTTACGCACCCAATTGGGTCCTCCTTCGGGGTTCAGGGCAA | 155215122 | g |
| chr1 | 155235918 | a | 155215236 | g |
| chr1 | 155236097 | g | 155215415 | c |
| chr1 | 155236102 | t | 155215420 | c |
| chr1 | 155236129 | t | 155215447 | c |
| chr1 | 155236145 | g | 155215463 | c |
| chr1 | 155236175 | g | 155215493 | a |
| chr1 | 155236190 | a | 155215508 | g |
| chr1 | 155237982 | t | 155216705 | c |

| | | | | |
|---|---|---|---|---|
| chr1 | 155237989 | g | 155216712 | a |
| chr1 | 155237990 | c | 155216713 | t |
| chr1 | 155237996 | g | 155216719 | t |
| chr1 | 155237997 | c | 155216720 | t |
| chr1 | 155238055 | t | 155216778 | c |
| chr1 | 155238088 | t | 155216811 | c |
| chr1 | 155238092 | t | 155216815 | g |
| chr1 | 155238141 | a | 155216864 | t |
| chr1 | 155238174 | c | 155216897 | t |
| chr1 | 155238192 | a | 155216915 | g |
| chr1 | 155238206 | a | 155216929 | c |
| chr1 | 155238214 | a | 155216937 | c |
| chr1 | 155238630 | g | 155217353 | a |
| chr1 | 155238754 | g | 155217477 | a |
| chr1 | 155238872 | ag | 155217595 | t |
| chr1 | 155238882 | a | 155217604 | g |
| chr1 | 155238929 | a | 155217651 | t |
| chr1 | 155238930 | g | 155217652 | t |

**Table S2:** *GBA* variants targeted by Gauchian. Coordinates are in hg38.

| chr | position | ref | alt | variant_name |
|---|---|---|---|---|
| chr1 | 155235002 | C | T | p.R535H |
| chr1 | 155235003 | G | A | p.R535C |
| chr1 | 155235007 | C | T | p.W533X |
| chr1 | 155235057 | C | T | p.G517S |
| chr1 | 155235101 | C | T | c.1506-1G>A |
| chr1 | 155235196 | G | A | p.R502C |
| chr1 | 155235197 | G | C | p.N501K |
| chr1 | 155235205 | C | G | p.V499L |
| chr1 | 155235217 | C | G | p.A495P |
| chr1 | 155235241 | C | T | p.A487T |
| chr1 | 155235252 | A | C | p.L483R |
| chr1 | 155235252 | A | G | p.L483P |
| chr1 | 155235303 | A | C | p.I466S |
| chr1 | 155235679 | A | G | c.1388+2T>C |

| | | | | |
|---|---|---|---|---|
| chr1 | 155235708 | G | C | p.P454R |
| chr1 | 155235712 | G | A | p.Q453X |
| chr1 | 155235721 | A | T | p.F450I |
| chr1 | 155235726 | T | A | p.D448V |
| chr1 | 155235727 | C | G | p.D448H |
| chr1 | 155235749 | GGGACTGTCGACAAAGTTACGCACCCAATTGGGTCCTCCTTCGGGGTTCAGGGCAA | G | p.L422fs/c.1263del |
| chr1 | 155235750 | G | A | p.P440L |
| chr1 | 155235757 | C | T | p.D438N |
| chr1 | 155235760 | C | A | p.V437F |
| chr1 | 155235772 | C | A | p.V433L |
| chr1 | 155235780 | G | A | p.P430L |
| chr1 | 155235790 | C | A | p.E427X |
| chr1 | 155235798 | A | G | p.L424P |
| chr1 | 155235810 | C | T | p.W420X |
| chr1 | 155235819 | C | T | p.W417X |
| chr1 | 155235823 | C | T | p.G416S |
| chr1 | 155235829 | C | A | p.V414L |
| chr1 | 155235831 | T | G | p.H413P |
| chr1 | 155235841 | G | C | p.L410V |
| chr1 | 155235843 | T | C | p.N409S |
| chr1 | 155235843 | T | G | p.N409T |
| chr1 | 155236255 | C | G | p.S405T |
| chr1 | 155236261 | C | G | p.S403T |
| chr1 | 155236277 | G | A | p.R398X |
| chr1 | 155236285 | G | A | p.S395F |
| chr1 | 155236292 | G | C | p.L393V |
| chr1 | 155236295 | G | A | p.R392W |
| chr1 | 155236295 | G | C | p.R392G |
| chr1 | 155236298 | C | G | p.V391L |
| chr1 | 155236318 | G | A | p.S384F |
| chr1 | 155236328 | A | C | p.C381G |
| chr1 | 155236379 | C | T | p.G364R |
| chr1 | 155236384 | G | A | p.T362I |
| chr1 | 155236416 | C | A | p.W351C |

| | | | | |
|---|---|---|---|---|
| chr1 | 155236417 | C | G | p.W351S |
| chr1 | 155236420 | T | C | p.H350R |
| chr1 | 155236426 | G | A | p.A348V |
| chr1 | 155236439 | CA | C | p.K342_Y343insX |
| chr1 | 155237099 | G | T | c.999+242C>A |
| chr1 | 155237357 | G | A | p.P328L |
| chr1 | 155237370 | G | A | p.R324C |
| chr1 | 155237394 | G | A | p.R316C |
| chr1 | 155237411 | C | T | p.S310N |
| chr1 | 155237412 | T | C | p.S310G |
| chr1 | 155237425 | AG | A | p.P305fs |
| chr1 | 155237427 | G | C | p.P305A |
| chr1 | 155237444 | A | G | p.I299T |
| chr1 | 155237446 | G | T | p.F298L |
| chr1 | 155237453 | C | T | p.R296Q |
| chr1 | 155237454 | G | A | p.R296X |
| chr1 | 155237470 | G | T | p.F290L |
| chr1 | 155237474 | C | G | p.G289A |
| chr1 | 155237480 | C | A | p.C287F |
| chr1 | 155237576 | A | T | p.F255Y |
| chr1 | 155237577 | A | C | p.F255V |
| chr1 | 155237579 | C | G | c.762-1G>C |
| chr1 | 155238141 | A | T | p.F252I |
| chr1 | 155238174 | C | T | p.G241R |
| chr1 | 155238186 | T | C | p.K237E |
| chr1 | 155238192 | A | G | p.S235P |
| chr1 | 155238194 | C | T | p.G234E |
| chr1 | 155238214 | A | C | p.N227K |
| chr1 | 155238214 | AT | CC | p.N227R |
| chr1 | 155238215 | T | C | p.N227S |
| chr1 | 155238228 | A | G | p.W223R |
| chr1 | 155238234 | G | T | p.P221T |
| chr1 | 155238242 | C | T | p.W218X |
| chr1 | 155238264 | CG | C | p.V211fs |

| | | | | |
|---|---|---|---|---|
| **chr1** | 155238270 | G | A | p.R209C |
| **chr1** | 155238298 | CAG | C | p.L199fs |
| **chr1** | 155238519 | T | G | p.K196Q |
| **chr1** | 155238525 | T | A | p.K194X |
| **chr1** | 155238547 | GA | G | p.F186fs |
| **chr1** | 155238571 | AG | A | p.P178fs |
| **chr1** | 155238579 | C | T | p.D176N |
| **chr1** | 155238584 | T | C | p.Y174C |
| **chr1** | 155238596 | C | A | p.R170L |
| **chr1** | 155238596 | C | T | p.R170H |
| **chr1** | 155238597 | G | A | p.R170C |
| **chr1** | 155238608 | T | A | p.D166V |
| **chr1** | 155238617 | GC | G | p.A163fs |
| **chr1** | 155238624 | G | A | p.P161S |
| **chr1** | 155238629 | C | T | p.R159Q |
| **chr1** | 155238630 | G | A | p.R159W |
| **chr1** | 155239639 | A | C | p.L144R |
| **chr1** | 155239655 | C | G | p.A139P |
| **chr1** | 155239665 | CAG | C | p.L135fs |
| **chr1** | 155239716 | C | G | p.K118N |
| **chr1** | 155239736 | G | A | p.Q112X |
| **chr1** | 155239934 | G | A | p.R87W |
| **chr1** | 155239939 | C | T | p.G85E |
| **chr1** | 155239968 | GGTA | G | p.T75del |
| **chr1** | 155239989 | C | CG | p.T69fs |
| **chr1** | 155240033 | C | A | p.V54L |
| **chr1** | 155240629 | C | T | c.115+1G>A |
| **chr1** | 155240637 | C | T | p.W36X |
| **chr1** | 155240651 | G | GC | p.Q32fs |
| **chr1** | 155240660 | G | GC | p.L29fs |
| **chr1** | 155240671 | AG | A | p.L25fs |
| **p.L483P + p.A495P + p.Val499=** | | | | RecNciI |
| **RecNciI + p.D448H** | | | | Rec.TL |
| **RecNciI + p.D448H + c.1263del** | | | | c.1263del+RecTL |

**Table S3:** PCR conditions for primer pair A.

| Reagents | Volume | | °C | Time | cycles |
|---|---|---|---|---|---|
| Template DNA (200ng) | x µL | | 94 | 30sec | 1 |
| Nuclease free water | 19µL - x | | 94 | 15sec | |
| Mg2+ | 2µL | | 65 | 15sec | 35 |
| Forward primer | 2µL | | 65 | 6min | |
| Reverse primer | 2µL | | 65 | 10min | 1 |
| LongAmp Taq 2x polymerase mix (NEB) | 25µL | | 4 | hold | |
| Primer pair A* | Forward | 5′- TTTCTGTTGGTGCTGATATTGCTCCTAAAGTTGTCACCCATACATG -3′ | | | |
| | Reverse | 5′- ACTTGCCTGTCGCTCTATCTTCCCAACCTTTCTTCCTTCTTCTCAA -3′ | | | |

*primers have been edited to contain the ONT barcode adapter sequence.

**Table S4:** PCR conditions for primers pairs B and C.

| Reagents | Volume | | °C | Time | cycles |
|---|---|---|---|---|---|
| Template DNA (100ng) | x µL | | 95 | 3min | 1 |
| Nuclease free water | 24.5µL - x | | 95 | 15sec | |
| | | | 62 | 15sec | 12 |
| Barcode mix | 0.5µL | | 65 | 10min | |
| | | | 65 | 10min | 1 |
| LongAmp Taq 2x polymerase mix (NEB) | 25µL | | 4 | hold | |
| Primer pair B* | Forward | 5′-TTTCTGTTGGTGCTGATATTGCATGTGTCCATTCTCCATGTCTTCA-3′ | | | |
| | Reverse | 5′-ACTTGCCTGTCGCTCTATCTTCAGCCTTCCTTCCTTCCCTGCAT-3′ | | | |
| Primer pair C* | Forward | 5′-TTTCTGTTGGTGCTGATATTGCGTGTCCGTTCTCCACATCCTTG-3′ | | | |
| | Reverse | 5′-ACTTGCCTGTCGCTCTATCTTCCCAACCTTTCTTCCTTCTTCTCAA-3′ | | | |

*primers have been edited to contain the ONT barcode adapter sequence.

**Table S5:** Results of dPCR. The mean dPCR copy number of 2 probes is given.

| Sample | Copy number detected by Gauchian | Copy number (dPCR) | ONT CNV detected |
|---|---|---|---|
| NA20756 | 3 | 3.18 | Duplication |
| HG01912 | 5 | 5.15 | Duplication |
| HG01889 | 7 | 6.71 | Duplication |
| HG02284 | 8 | 7.86 | Duplication |
| RAP1 | not analysed | 0.98 | Fusion |
| RAP2 | not analysed | 2.06 | None |

**Table S6:** Phenotypes and genotypes of all samples from the RAPSODI cohort.

| | Wild type | Heterozygous carriers* | Homozygous and compound heterozygous carriers* | Total |
|---|---|---|---|---|
| PD Patients | 195 | 36 | 0 | 231 |
| GD Patients | 0 | 0 | 33 | 33 |
| Healthy individuals | 74 | 43 | 0 | 117 |
| total | 269 | 79 | 33 | 381 |

*Including pathogenic structural variants.

**Table S7:** All variants detected in the RAPSODI and QSBB cohorts.

| Allele detected | Number of alleles |
|---|---|
| p.N409S (NC_000001.11:g.155235843T>G) | 35 |
| p.L483P (NC_000001.11:g.155235252A>G) | 19 |
| p.E365K (NC_000001.11:g.155236376C>T) | 15 |
| p.T408M (NC_000001.11:g.155236246G>A) | 4 |
| p.R502C (NC_000001.11:g.155235196G>A) | 3 |
| IVS6-2 (NC_000001.11:155238308T>C) | 2 |
| p.R301H (NC_000001.11:g.155237438C>T) | 2 |
| p.84GG (NC_000001.11:g.155240661dup) | 1 |
| p.D419N (NC_000001.11:g.155235814C>T) | 1 |
| IVS9+1 (NC_000001.11:g.155235680C>T) | 1 |
| p.K13R (NC_000001.11:g.155240707T>C) | 1 |
| p.R301G (NC_000001.11:g.155237439G>C) | 1 |
| p.T270I (NC_000001.11:g.155237531G>C) | 1 |
| p.T408M + L104= (NC_000001.11:g.155236246G>A + 155239758C>T) | 1 |
| p.A357D (155236399G>T) | 1 |
| p.D354H (NC_000001.11:g.155236409C>G) | 1 |
| p.G241R (NC_000001.11:g.155238174C>T) | 1 |
| p.L519P (NC_000001.11:g.155235050A>G) | 1 |
| p.P211T (NC_000001.11:g.155238234G>T) | 1 |
| c.413delC (NC_000001.11:g.155239661del) | 1 |
| RecNciI(p.A495P + p.Val499= + p.L483P) / reciprocal | 5 |
| RecNciI(p.A495P + p.Val499= + p.L483P) / non-reciprocal | 1 |
| RecTL (p.A495P + p.Val499= + p.L483P + p.D448H) + c.1263del / non-reciprocal | 2 |
| Non-pathogenic reciprocal CN loss recombinants | 4* |
| Non-pathogenic reciprocal CN gain recombinants | 7** |

These do not include the variants found in the 95 samples previously reported (Salazar et al, 2019)
*In two alleles, a non-pathogenic CNL was detected in cis with p.L444P
**In two samples, a CNG was detected together with c.1263del+RecTL (gene conversion). In two other samples, a duplication recombinant was detected together with the p.L483P variant. These were not phased.

**Table S8:** List of *GBAP1*-like variant calls in Exon 9-11 homology region in 1kGP and AMP-PD cohorts.

| sampleID | IsBiallelic | IsCarrier | CN(GBA+ GBAP1) | Deletion breakpoint in *GBA* | Recombinant variant allele1 | Recombinant variant allele2 |
|---|---|---|---|---|---|---|
| HG00422 | FALSE | TRUE | 3 | TRUE | RecNciI | |
| LB-00540 | FALSE | TRUE | 3 | TRUE | RecNciI | |
| LB-00542 | FALSE | TRUE | 3 | TRUE | RecNciI | |
| LB-00599 | FALSE | TRUE | 3 | TRUE | RecNciI | |
| LB-00695 | FALSE | TRUE | 3 | TRUE | RecNciI | |
| LB-01632 | FALSE | TRUE | 3 | TRUE | c.1263del+RecTL | |
| LB-01652 | FALSE | TRUE | 3 | FALSE | L483P | |
| LB-01755 | FALSE | TRUE | 3 | TRUE | RecNciI | |
| LB-01782 | FALSE | TRUE | 3 | TRUE | RecNciI | |
| LB-01788 | FALSE | TRUE | 3 | TRUE | RecNciI | |
| LB-02286 | FALSE | TRUE | 3 | TRUE | RecNciI | |
| LB-02338 | FALSE | TRUE | 3 | TRUE | RecNciI | |
| LB-02662 | FALSE | TRUE | 3 | TRUE | c.1263del+RecTL | |
| LB-02817 | FALSE | TRUE | 3 | TRUE | RecNciI | |
| HB-PD_INVDX373MWV | FALSE | TRUE | 3 | FALSE | L483P | |
| HB-PD_INVWN635UXR | FALSE | TRUE | 3 | TRUE | RecNciI | |
| PD-PDEK306DBC | FALSE | TRUE | 3 | FALSE | L483P | |
| PD-PDLR687WTH | FALSE | TRUE | 3 | TRUE | c.1263del+RecTL | |
| HG00119 | FALSE | TRUE | 4 | None | c.1263del+RecTL | |
| NA20815 | FALSE | TRUE | 4 | None | L483P | |
| NA18991 | FALSE | TRUE | 4 | None | L483P | |
| HG02613 | FALSE | TRUE | 4 | None | A495P | |
| HG00115 | FALSE | TRUE | 4 | None | c.1263del+RecTL | |
| HG02439 | FALSE | TRUE | 4 | None | c.1263del | |
| NA20513 | FALSE | TRUE | 4 | None | L483P | |
| HG00310 | FALSE | TRUE | 4 | None | L483P | |
| HG02151 | FALSE | TRUE | 4 | None | c.1263del | |
| HG03967 | FALSE | TRUE | 4 | None | L483P | |
| LB-00009 | FALSE | TRUE | 4 | None | RecNciI | |
| LB-00092 | FALSE | TRUE | 4 | None | L483P | |
| LB-00483 | FALSE | TRUE | 4 | None | RecNciI | |
| LB-00490 | FALSE | TRUE | 4 | None | L483P | |
| LB-00575 | FALSE | TRUE | 4 | None | c.1263del | |
| LB-00668 | FALSE | TRUE | 4 | None | L483P | |
| LB-00734 | FALSE | TRUE | 4 | None | L483P | |
| LB-00760 | FALSE | TRUE | 4 | None | L483P | |
| LB-00906 | FALSE | TRUE | 4 | None | A495P | |

| | | | | | |
|---|---|---|---|---|---|
| LB-01188 | FALSE | TRUE | 4 | None | L483P |
| LB-01223 | FALSE | TRUE | 4 | None | c.1263del |
| LB-01297 | FALSE | TRUE | 4 | None | L483P |
| LB-01339 | FALSE | TRUE | 4 | None | A495P |
| LB-01351 | FALSE | TRUE | 4 | None | L483P |
| LB-01363 | FALSE | TRUE | 4 | None | L483P |
| LB-01388 | FALSE | TRUE | 4 | None | L483P |
| LB-01391 | FALSE | TRUE | 4 | None | L483P |
| LB-01403 | FALSE | TRUE | 4 | None | L483P |
| LB-01440 | FALSE | TRUE | 4 | None | A495P |
| LB-01477 | FALSE | TRUE | 4 | None | c.1263del |
| LB-01540 | FALSE | TRUE | 4 | None | D448H |
| LB-01599 | FALSE | TRUE | 4 | None | L483P |
| LB-01613 | FALSE | TRUE | 4 | None | D448H |
| LB-01631 | FALSE | TRUE | 4 | None | RecNcil |
| LB-01696 | FALSE | TRUE | 4 | None | L483P |
| LB-01752 | FALSE | TRUE | 4 | None | L483P |
| LB-01803 | FALSE | TRUE | 4 | None | L483P |
| LB-01894 | FALSE | TRUE | 4 | None | L483P |
| LB-01922 | FALSE | TRUE | 4 | None | L483P |
| LB-02197 | FALSE | TRUE | 4 | None | L483P |
| LB-02483 | FALSE | TRUE | 4 | None | A495P |
| LB-02676 | FALSE | TRUE | 4 | None | c.1263del |
| LB-02760 | FALSE | TRUE | 4 | None | c.1263del |
| LB-02784 | FALSE | TRUE | 4 | None | L483P |
| LB-02815 | FALSE | TRUE | 4 | None | c.1263del+RecTL |
| LB-08162 | FALSE | TRUE | 4 | None | c.1263del |
| LB-06559 | FALSE | TRUE | 4 | None | A495P |
| LB-07084 | FALSE | TRUE | 4 | None | D448H |
| LB-07658 | FALSE | TRUE | 4 | None | A495P |
| BF-1080 | FALSE | TRUE | 4 | None | A495P |
| HB-PD_INVHN100ZHK | FALSE | TRUE | 4 | None | A495P |
| HB-PD_INVKJ120VLF | FALSE | TRUE | 4 | None | L483P |
| HB-PD_INVWU757PGL | FALSE | TRUE | 4 | None | RecNcil |
| PD-PDCH154ZHF | FALSE | TRUE | 4 | None | D448H |
| PD-PDDJ206TP0 | FALSE | TRUE | 4 | None | RecNcil |
| PD-PDEW749KZU | FALSE | TRUE | 4 | None | L483P |
| PD-PDMD724EA9 | FALSE | TRUE | 4 | None | L483P |

| | | | | | | |
|---|---|---|---|---|---|---|
| PD-PDXU108EDQ | FALSE | TRUE | 4 | None | L483P | |
| PD-PDXU630RLV | FALSE | TRUE | 4 | None | L483P | |
| PP-3420 | FALSE | TRUE | 4 | None | L483P | |
| PP-3429 | FALSE | TRUE | 4 | None | A495P | |
| PP-3700 | FALSE | TRUE | 4 | None | L483P | |
| PP-41342 | FALSE | TRUE | 4 | None | L483P | |
| PP-60060 | FALSE | TRUE | 4 | None | L483P | |
| HB-PD_INVAV028JED | FALSE | TRUE | 4 | None | L483P | |
| HB-PD_INVLR599UE9 | FALSE | TRUE | 4 | None | D448H | |
| HB-PD_INVLU007ANL | FALSE | TRUE | 4 | None | L483P | |
| PD-PDGF687UAV | FALSE | TRUE | 4 | None | L483P | |
| PP-57787 | FALSE | TRUE | 4 | None | L483P | |
| PP-59343 | FALSE | TRUE | 4 | None | L483P | |
| PP-59926 | FALSE | TRUE | 4 | None | L483P | |
| LB-00239 | FALSE | TRUE | 5 | None | L483P | |
| BF-1016 | FALSE | TRUE | 5 | None | L483P | |
| HB-PD_INVRH572AT6 | FALSE | TRUE | 5 | None | L483P | |
| PP-3307 | FALSE | TRUE | 5 | None | L483P | |
| LB-00823 | FALSE | TRUE | 6 | None | D448H | |
| LB-02672 | TRUE | FALSE | 7 | None | D448H | L483P |
| LB-01354 | FALSE | TRUE | 8 | None | c.1263del+RecTL | |

**Table S9:** Complete list of the number of samples carrying *GBA* variants in population samples.

| | 1kGP | PD | | LBD | | |
|---|---|---|---|---|---|---|
| **Variant** | N=2405 | case N=2325 | control N=1255 | case N=2598 | control N=1941 | mild/severe |
| **c.115+1G>A** | 2 | 3 | 1 | 3 | 0 | severe |
| **c.1263del** | 2 | 0 | 0 | 6 | 0 | severe |
| **c.1263del+RecTL** | 2 | 1 | 0 | 4 | 0 | severe |
| **p.A348V** | 0 | 0 | 0 | 1 | 0 | mild |
| **p.A495P** | 1 | 3 | 0 | 4 | 2 | mild |
| **p.D438N** | 0 | 0 | 0 | 1 | 0 | severe |
| **p.D448H** | 0 | 1 | 1 | 4 | 1 | severe |
| **p.E365K** | 25 | 91 | 21 | 145 | 30 | non-pathogenic (PD risk) |
| **p.F252I** | 0 | 1 | 0 | 0 | 0 | severe |
| **p.F298L** | 0 | 0 | 0 | 1 | 0 | severe |
| **p.G241R** | 0 | 1 | 0 | 4 | 0 | severe |
| **p.G364R** | 0 | 0 | 0 | 1 | 0 | severe |
| **p.G416S** | 0 | 0 | 0 | 1 | 0 | mild |
| **p.H350R** | 0 | 0 | 0 | 1 | 0 | severe |
| **p.K118N** | 0 | 0 | 0 | 1 | 0 | mild |
| **p.L29fs** | 0 | 3 | 0 | 0 | 1 | severe |
| **p.L483P** | 5 | 14 | 6 | 23 | 0 | severe |
| **p.L483R** | 0 | 1 | 0 | 0 | 0 | severe |
| **p.N227S** | 1 | 0 | 0 | 1 | 0 | mild |
| **p.N409S** | 3 | 128 | 158 | 59 | 19 | mild |
| **p.P305fs** | 0 | 0 | 0 | 1 | 0 | severe |
| **p.R159W** | 0 | 3 | 0 | 3 | 0 | severe |
| **p.R170C** | 0 | 0 | 0 | 1 | 0 | severe |
| **p.R296Q** | 0 | 3 | 1 | 3 | 0 | severe |
| **p.R398X** | 0 | 0 | 0 | 1 | 0 | severe |
| **p.R502C** | 2 | 2 | 0 | 8 | 0 | severe |
| **p.R535C** | 0 | 0 | 0 | 1 | 0 | mild |
| **p.R535H** | 0 | 6 | 3 | 4 | 1 | mild |
| **p.R87W** | 0 | 0 | 0 | 1 | 0 | mild |
| **p.S235P** | 0 | 1 | 0 | 0 | 0 | severe |
| **p.T408M** | 9 | 38 | 10 | 65 | 32 | non-pathogenic (PD risk) |
| **p.V433L** | 0 | 1 | 0 | 1 | 0 | severe |
| **p.V499L** | 0 | 0 | 0 | 1 | 0 | mild |
| **p.W223R** | 0 | 0 | 0 | 1 | 0 | severe |
| **p.W420X** | 0 | 0 | 0 | 1 | 0 | unknown |
| **RecNciI** | 1 | 3 | 0 | 13 | 0 | severe |
